# Stratifying Risk for Postpartum Depression at Time of Hospital Discharge

**DOI:** 10.1101/2024.05.27.24307973

**Authors:** Mark A. Clapp, Victor M. Castro, Pilar Verhaak, Thomas H. McCoy, Lydia L. Shook, Andrea G. Edlow, Roy H. Perlis

## Abstract

**Objective:** Postpartum depression (PPD) represents a major contributor to postpartum morbidity and mortality. Beyond efforts at routine screening, risk stratification models could enable more targeted interventions in settings with limited resources. Thus, we aimed to develop and estimate the performance of a generalizable risk stratification model for PPD in patients without a history of depression using information collected as part of routine clinical care.

**Methods:** We performed a retrospective cohort study of all individuals who delivered between 2017 and 2022 in one of two large academic medical centers and six community hospitals. An elastic net model was constructed and externally validated to predict PPD using sociodemographic factors, medical history, and prenatal depression screening information, all of which was known before discharge from the delivery hospitalization.

**Results:** The cohort included 29,168 individuals; 2,703 (9.3%) met at least one criterion for postpartum depression in the 6 months following delivery. In the external validation data, the model had good discrimination and remained well-calibrated: area under the receiver operating characteristic curve 0.721 (95% CI: 0.707-0.734), Brier calibration score 0.088 (95% CI: 0.084 – 0.092). At a specificity of 90%, the positive predictive value was 28.0% (95% CI: 26.0-30.1%), and the negative predictive value was 92.2% (95% CI: 91.8-92.7%).

**Conclusions:** These findings demonstrate that a simple machine-learning model can be used to stratify the risk for PPD before delivery hospitalization discharge. This tool could help identify patients within a practice at the highest risk and facilitate individualized postpartum care planning regarding the prevention of, screening for, and management of PPD at the start of the postpartum period and potentially the onset of symptoms.

## Introduction

Postpartum depression (PPD) is common, affecting approximately 15% of recently pregnant individuals, and represents a major contributor to both morbidity and mortality following pregnancy.^1–6^ It is associated with an increased risk for suicide and self-harm and is estimated to be responsible for 10% or more of all pregnancy-related deaths.^7–9^ Compared to other causes of pregnancy-related deaths, those due to mental health conditions are considered preventable by many Maternal Mortality Review Committees.^7^ In addition to the increased risk of self-harm, PPD also has a profound impact on a person’s physical and mental health, ability to function, and relationships with their newborn and family during a period that can represent a challenge even in the absence of mood change.^2,10^

As >98% of women deliver in a hospital or health care facility, the delivery hospitalization represents an opportunity to identify individuals at high risk for postpartum depression and potentially target interventions, prevent, screen, and manage PPD.^11^ Such interventions could be applied more efficiently if risk could more readily be determined; for this reason, scales like the Edinburgh Postpartum Depression Scale (EPDS) are increasingly incorporated into obstetric care.^12,13^ However, while current symptoms are useful for prediction, the ability of the EPDS to characterize risk remains modest.^14^

To date, most attempts to develop a usable PPD risk stratification tool in practice have been limited because they lack external validation, such that estimates of performance are likely optimistic, or they include individuals who are currently depressed or treated, which is likely to yield inflated estimates of performance as individuals who are depressed before delivery are substantially more likely to be depressed after delivery.^15–17^ To address this gap, we sought to develop and externally validate a simple, machine-learning-based PPD risk model using electronic health record (EHR) information known by the clinical team during hospitalization for delivery for individuals with no recent history of a depressive disorder, which could be used by better stratify risk and target prevention strategies or resources to support recently postpartum individuals.

## Methods

### Study Design and Data

We examined all live births between 2017 and 2022 in coded clinical data in 2 academic medical centers, 6 community-based hospitals, and their affiliated outpatient clinics, all sharing a common electronic health record (EHR). Patients from these hospitals were split into 2 groups with roughly equal sizes: the model development group contained all patients delivering at 5 hospitals, including 1 of the academic medical centers, and the model validation group contained all patients delivering at the other 3 hospitals, including 1 of the academic medical centers. For the primary study cohort, we excluded individuals who did not receive routine prenatal care at any of the included hospitals or networks, as these predominantly reflect individuals treated in practices that do not use the same electronic health record system as the delivery hospitals (e.g., private practices) because their medical history could not be observed. We further excluded individuals with a diagnostic code reflecting mood or psychotic disorder (Supplemental Table S1) or who had an antidepressant prescription (Supplemental Table S12) in the 12 months preceding delivery, as these individuals would be considered high risk for PPD based on their prior clinical history and/or already receiving mental health care.

The Massachusetts General Brigham Institutional Review Board approved this study. We followed the TRIPOD reporting guideline for model development and validation.^18^ All analyses used R 4.2.2 (The R Foundation for Statistical Computing, Vienna, Austria). To facilitate interpretation, we present 95% confidence intervals for all estimates rather than p-values.

### Predictor Variables

We considered information that would be known by the clinical team at the time of delivery hospitalization as predictor variables for the model, including maternal medical history, medication use, pregnancy history, and demographic factors. The medical history of the birthing person was captured in terms of ICD-10 diagnosis codes that occurred within 1 year before the admission date for the delivery admission. Codes were grouped using the Agency for Healthcare Research and Quality’s Clinical Classification Software – Refined (CCSR)^19^; codes that occurred in less than 20 individuals were excluded. Similarly, all medication data in the EHR listed or present within 1 year before the admission date for the delivery admission were were grouped at both the RxNorm drug ingredient level and by drug class^20^; medications that observed in less than 20 individuals were excluded. We also incorporated data elements known or hypothesized to influence the risk of PPD, including maternal age at delivery, self-reported sex at birth, education (college degree vs. no college degree), marital status (single vs. not single), self-reported primary language (English vs. non-English), insurance type (public vs. private), and pregnancy factors: gestational age at delivery (term vs. preterm), number of gestations (singleton vs. multiple gestations), mode of delivery (vaginal vs. cesarean), number of prenatal visits, and delivery admission length of stay. Missing data for categorical data were included in the model as a unique group; missing data for pre-pregnancy BMI was replaced with the training set mean.. We also extracted self-reported race and ethnicity; these variables were not used for modeling but for subsequent subgroup analyses.

The American College of Obstetricians and Gynecologists (ACOG) recommends that all postpartum individuals receive perinatal depression screening at a postpartum visit^21^. However, ACOG’s guidelines for “Screening for Perinatal Depression” does also encourage, but not explicitly recommend, depression screening during pregnancy before the postpartum period. In our preliminary review, we observed that EPDS was administered prenatally in approximately 40% of patients who delivered at a hospital within the health system, which was highly dependent on local practice patterns. As we hypothesized that prenatal EPDS would potentially be strongly related to the risk of PPD, our primary model was restricted to individuals who had prenatal EPDS recorded. For individuals who had multiple prenatal EPDS values recorded during the pregnancy, the highest score was used.

### Outcome Definition

The primary outcome was PPD defined by the presence of either a mood or psychotic disorder diagnostic code (Supplemental Table S1), an antidepressant prescription (Supplemental Table S2), or a positive screen on the postpartum EPDS (EPDS>=13) within 6 months of delivery.^22^ Sensitivity analysis examined these outcomes individually. While postpartum psychosis may represent a distinct phenotype, we did not exclude it from the definition of PPD given the rarity of this outcome and the clinical imperative to identify high-risk individuals.^23^

### Model Development and Evaluation

The deliveries at 1 academic medical center and 4 affiliated community hospitals (the model development group) were divided into a model training (75%) and testing (25%) set, with random assignment stratified by the component of the outcome composite, ensure an equivalent representation of PPD in each set. We used an elastic net model with grid search and optimized model parameters in the training set, ensuring that only the model development data were used for optimization. The primary model incorporated the prenatal EPDS score, sociodemographic features (other than race and ethnicity), medical history, medication data known before delivery, and delivery features. For both the training and testing sets, model discrimination was assessed by the area under the receiver operating characteristic curve (AUROC), as well as positive and negative predictive values (PPV, NPV) using a screen-positive threshold using a set specificity of 90%. Similarly, calibration was assessed by Brier class score and by comparing observed, expected event rates by risk decile using calibration plots.

We next examined the model’s generalizability by evaluating discrimination and calibration in patients delivering at 3 separate hospitals (external validation group). Finally, we compared the primary model’s statistical parity in this external validation set among the subgroups of patients’ self-reported race, ethnicity, and age at delivery (categorized as less than 30, 30-40, and greater than 40 years) to evaluate if the model performed varied in different groups of people. This understanding can help, inform if there may be differential impacts by applying a universal, population-level screen-positive threshold among certain groups within a larger cohort.

Because EPDS was not universally collected or reported prenatally for all patients within the health system, we constructed, internally validated, and externally validated the same model without prenatal EPDS score in the same cohort in which the primary model was developed (i.e., prenatal EPDS known). Model discrimination and calibration were assessed and compared with the primary model that included EPDS.

## Results

Of the 98,620 deliveries in the full cohort, 80,027 (81.1%) received prenatal care within the networks affiliated with these hospitals, and 34,815 (35.3%) had an EPDS score recorded before the delivery encounter. Of the 29,168 with no prior diagnoses of depressive disorders or prescription for an antidepressant in the year prior to delivery, 15,018 were included in the model derivation group (51.5%), and 14,150 were in the model validation group (48.5%). The CONSORT diagram is shown in Figure 1.

**Figure 1.**
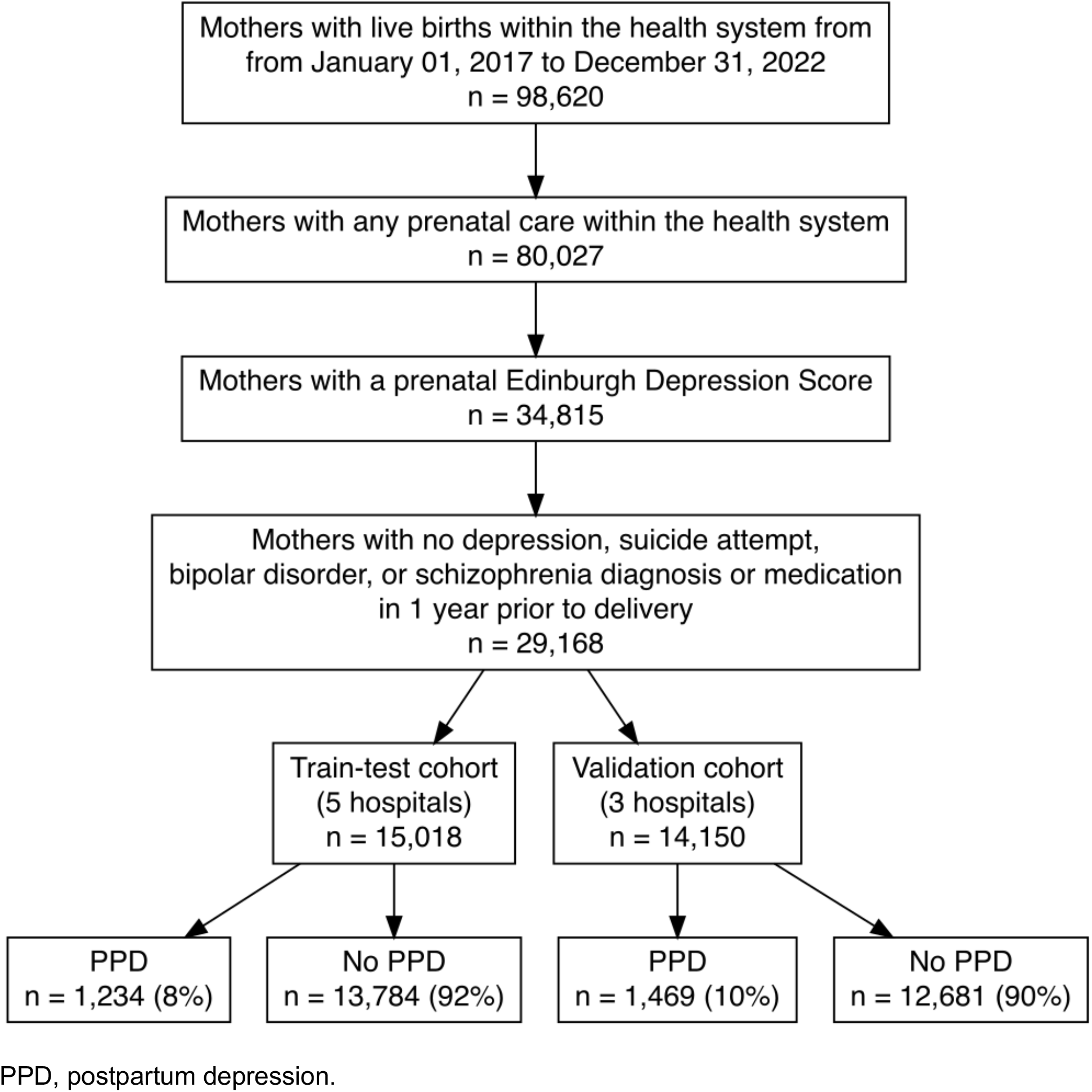
CONSORT Diagram

Table 1 summarizes the demographic and clinical characteristics of these individuals among the model development and external validation cohorts. Among all patients, the median age was 33 years [interquartile range (IQR) 30-36]. 3,675 (13%) self-identified as Asian, 1,996 (7%) as Black, 20,092 (70%) as White, 480 (1.7%) as >1 race, and 2,391 (8.4%) as another race not otherwise classified; 3,163 (11%) individuals were of Hispanic ethnicity. In the model derivation group, 1,234 individuals (8.2%) experienced at least one of the PPD outcomes, including 730 (4.9%) defined by diagnosis, 691 (4.6%) by medication, and 304 (2.0%) by EPDS. In the model validation group, 1,469 individuals (10.4%) experienced at least one of the PPD outcomes, including 869 (6.1%) defined by diagnosis, 826 (5.8%) by medication, and 388 (2.7%) by EPDS. For comparison, sociodemographic and clinical characteristics of those who developed and did not develop PPD are included in the Supplement (Table S3).

**Table 1.**
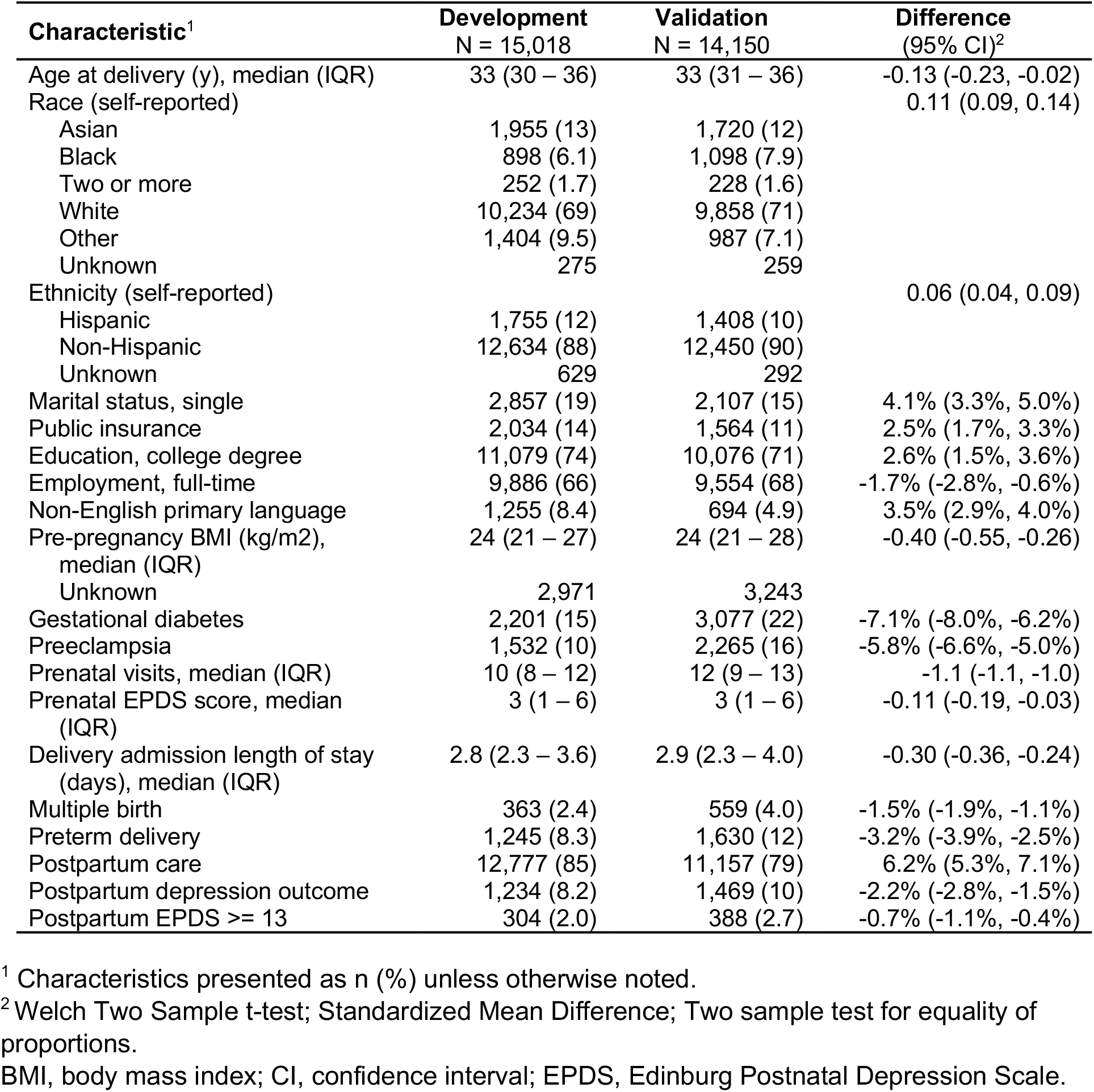
Sociodemographic and clinical characteristics of individuals in the model development (testing and training) and external validation data sets.

The ROC curve for the primary model is shown in Figure 2A for the testing data set in the model derivation group; in the same figure, the ROC curves for the models only with sociodemographic features without the prenatal EPDS score are also shown. The primary model features are summarized in Supplemental Figure 1. For the primary model, the AUROC was 0.736 (standard error (SE) 0.01), indicating the model had good discrimination, and the Brier score was 0.069 (SE 0.002]), indicating the model was well calibrated. At a set threshold of 90% specificity, the PPV was 24.2% and NPV was 93.7%. Figure 3A also illustrates calibration for this model by plotting the observed versus expected rates of PPD by risk decile.

**Figure 2.**
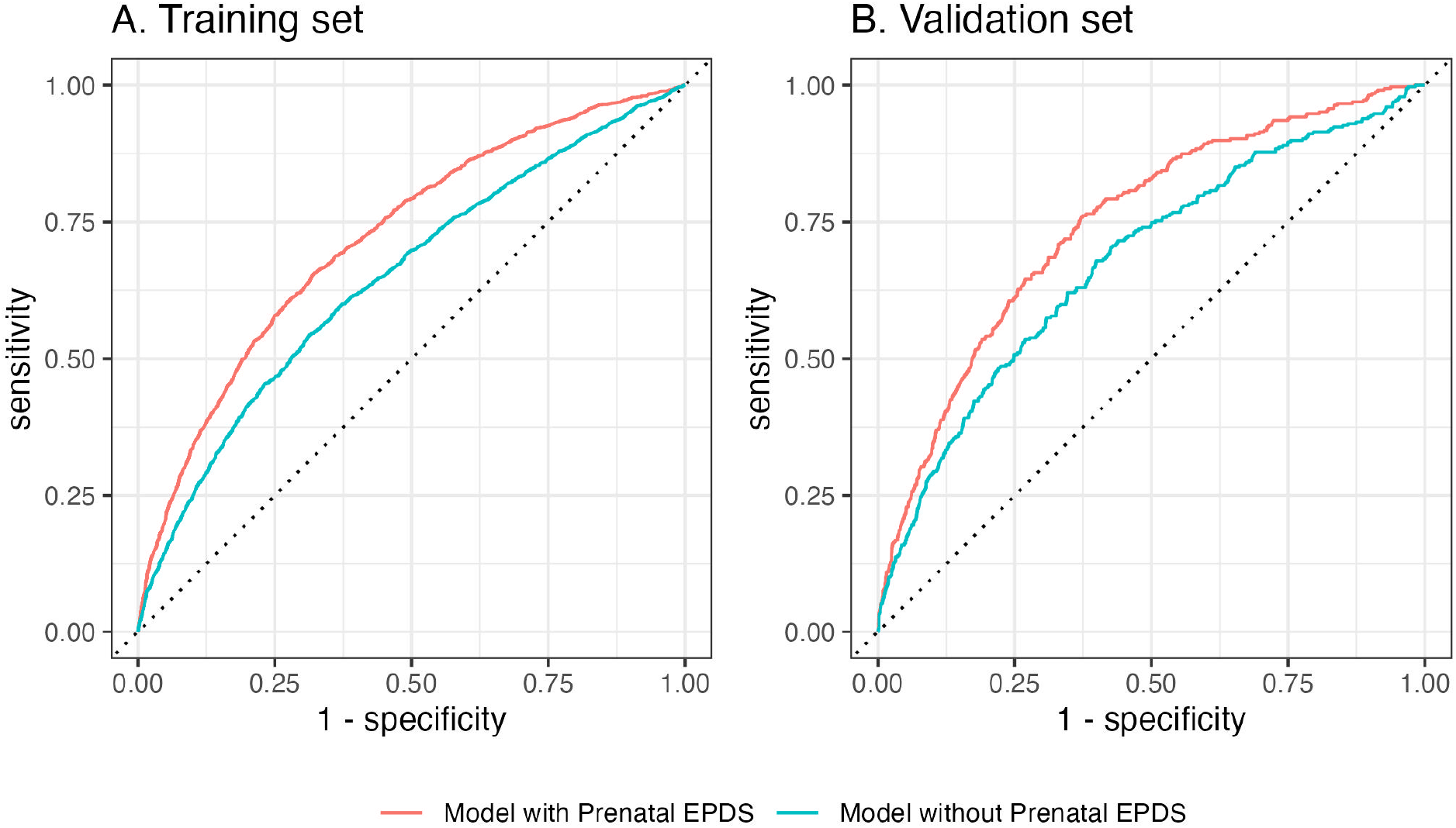
Postpartum depression (PPD) prediction model discrimination receiver operating characteristic curve in a random test (A) and independent external validation (B) cohort. The red line corresponds to the full elastic net model incorporating sociodemographic characteristics and prenatal diagnosis, medications, and Edinburgh postnatal depression score (EPDS). The blue line shows an elastic net model that includes all the same terms except prenatal EPDS.

**Figure 3.**
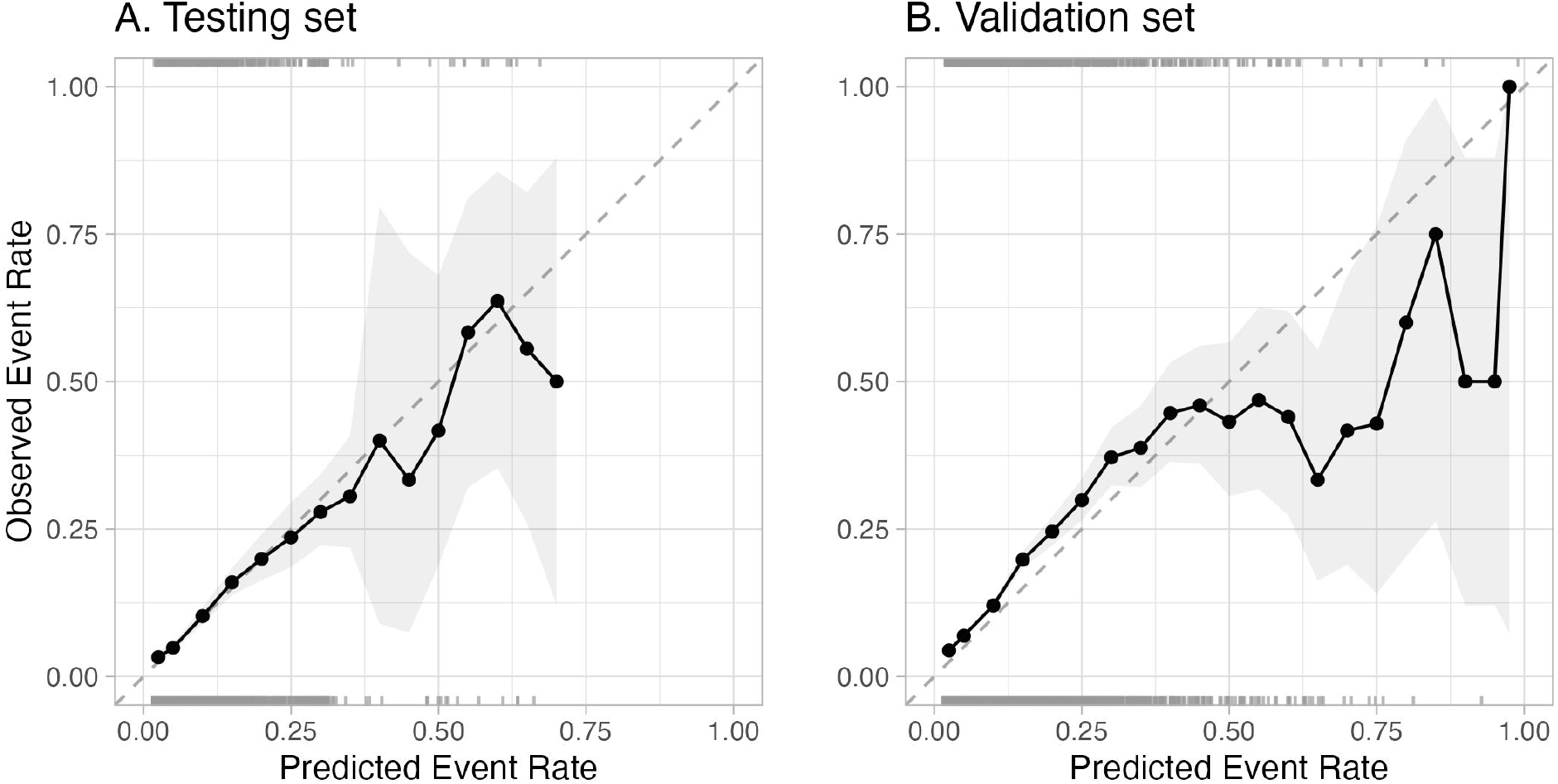
Primary postpartum depression (PPD) prediction model calibration in a random test (A) and independent external validation (B) cohort. The line indicates the PPD rate at intervals of the model prediction score. The shaded area represents a 95% confidence interval.

To evaluate the model’s external validity, the ROC curve is shown in Figure 2B. In this separate cohort, the model performed similarly: AUROC 0.721 (95% CI: 0.707-0.734), Brier score 0.088 (95% CI: 0.084-0.092). At a specificity of 90%, the PPV was 28.0% (95% CI: 26.0-30.1%) and NPV was 92.2% (95% CI: 91.8%-92.7%). The calibration plot in the external validation data after logistic calibration is shown in Figure 3B.

As prenatal EPDS was not universally available within the cohort, we compared the primary model to one built from the same factors except for the prenatal EPDS score. The ROC curves are shown for comparison in the testing and validation sets in Figures 2A and 2B, respectively. For the external validation, the AUROC was 0.647 (95% CI: 0.632-0.662), and the Brier score was 0.091 (95% CI: 0.087-0.095). The calibration plots are shown in Figure 4. The discrimination was higher in the primary model, demonstrating how regular EPDS screening in the prenatal period could be used to better stratify risk of PPD in combination with other patient factors.

**Figure 4.**
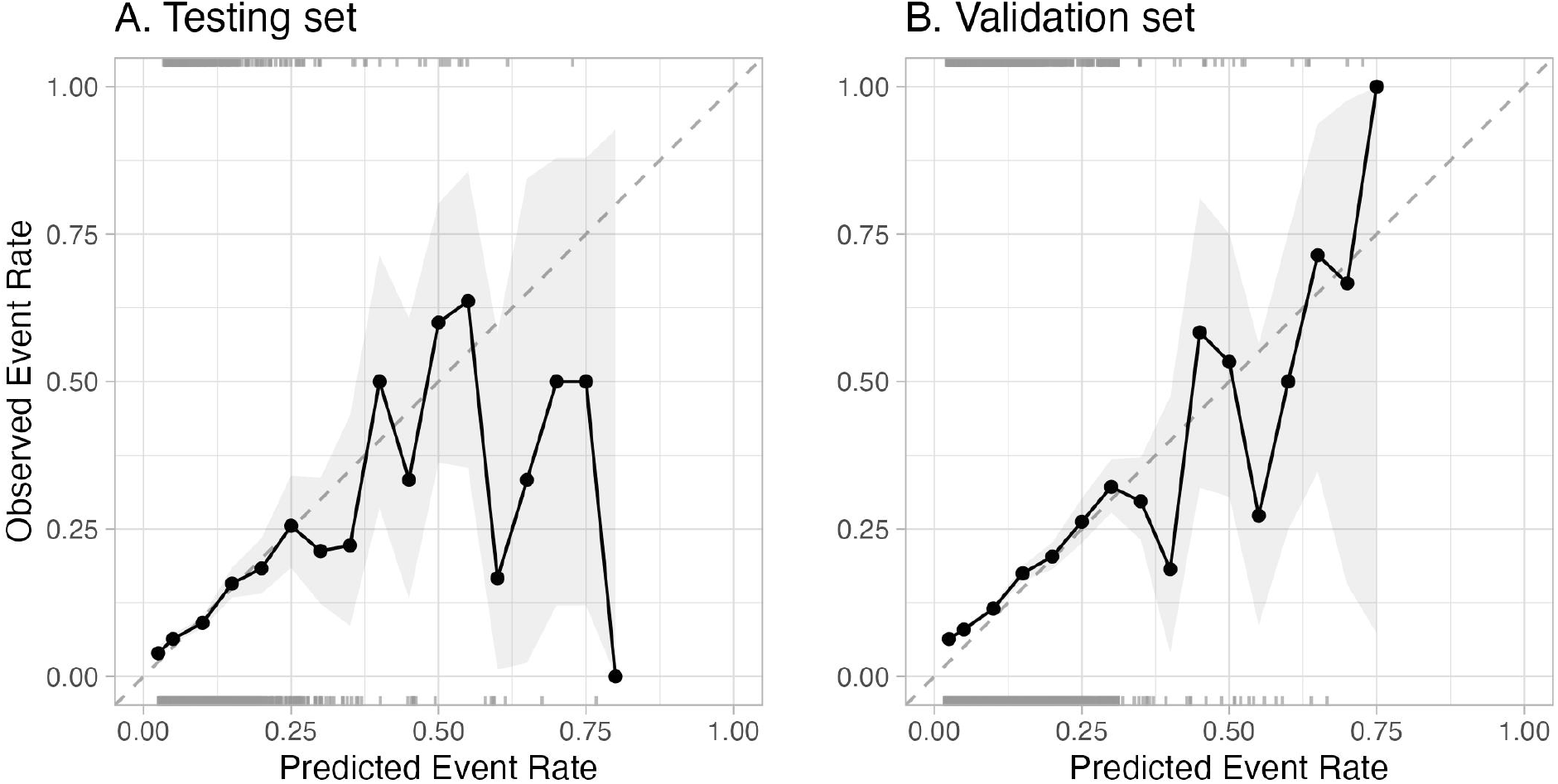
Postpartum depression (PPD) prediction model (excluding prenatal Edinburgh Postnatal Depression Scale score) calibration in a random test (A) and independent external validation (B) cohort. The line indicates the PPD rate at intervals of the model prediction score. The shaded area represents a 95% confidence interval.

Finally, we characterized fairness by examining model discrimination (AUROC) and calibration (Brier score) by subgroups of race, ethnicity, age, and hospital type (academic vs. community-based) in the validation set. Results are included in Supplemental Table S4. In general, the model performed similarly in all groups.

## Discussion

In this investigation of more than 29,000 deliveries without prior depression history across a large health system, the incidence of PPD was approximately 10%. A simple regression-based, machine-learning model achieved discrimination of 0.721 in the validation set. At a specificity of 90%, the PPV was 28%, nearly 3x the baseline population risk for PPD, and NPV was 92%. The model demonstrated reasonable calibration, distinguishing between higher and lower-risk populations. The risk model also had similar performance across patient subgroups, suggesting it could equitably be applied in a diverse population. These findings suggest that this model, using information known at the time of the delivery admission, could help clinical care teams stratify risk for PPD and assist in directing resources and support services to prevent and treat PPD, thereby reducing the subsequent morbidity of this relatively common condition.

Validation studies of the EPDS have demonstrated this tool’s ability to screen for depression during pregnancy and in the postpartum period^22^. However, the prenatal EPDS score has not been studied as a predictive tool for the development of PPD. We hypothesized prenatal EPDS would be an important feature in a PPD risk stratification model and thus limited our study population to those with an available score. When comparing the same models with and without the prenatal EPDS score, the model with the score had higher discrimination (AUC 0.72 vs. 0.65) in the external validation set. These findings suggest there may be multiple benefits to administering the EPDS screening tool prenatally, including screening for depression during pregnancy and also as a component of PPD risk stratification at the end of pregnancy.

Prior studies have attempted to stratify the risk of PPD but have limited utility or have not been externally validated; we contrast our model with those presented in three previously published studies. Wang et al. used EHR data to stratify the risk for PPD using coded data from 9,980 pregnancies and achieved an AUC of 0.79; however, this model was not externally validated.^16^ As with most subsequent studies, this model included individuals who had a pre-existing diagnosis of depression, which contributed to this model’s discriminatory performance.^16^ Wakefield and Frasch conducted a secondary analysis of the NIH *Nulliparous Pregnancy Outcomes Study: Monitoring Mothers-to-B*e study; using data from 10,000 individuals enrolled in this prospective investigation, they developed a model with excellent discrimination (AUC 0.91)^17^. However, this study had a very narrow subgroup of the population (i.e., nulliparous patients who agreed to be prospectively followed in a clinical trial) and included individuals who could have current or recent depression, with these features explaining much of the variance in the resulting models (i.e., individuals depressed before delivery are likely, continue, be depressed)^17^. One of the largest studies to date that developed and externally validated a prediction model for PPD used Danish registry data.^15^ This ambitious effort yielded very promising AUCs (>0.80) but reflected a model constructed from data from a national health system registry and excluded individuals with any prior psychiatric contact, diminishing transferability to US health systems and general clinical populations. In contrast, our study represents a more generalizable approach, predicting PPD among individuals not already known to be high risk based on a prior history of depression in a diverse US-based cohort using information that would be known by care teams before discharge from the delivery hospitalization.

Our study did not examine how and when this information could be presented to care team members or patients or used in practice. This is an important next step of this work, as we have shown that certain presentations of risk information can adversely influence provider behavior and decision-making.^24^ Similarly, a recent study examined multiple strategies for presenting postpartum depression risk to patients; the study showed that while the information while could be conveyed accurately, the format of the information influenced patient trust, behavior, and risk perception.^25^ Future work is planned to study the impact of implementing this risk stratification system in practice on the screening, prevention strategies, and management of PPD, which affects over 10% of postpartum individuals.

This study has several limitations. First, as with any study using diagnosis codes, there is a possibility of misclassification. The effects of missed or inaccurate diagnoses are difficult to predict; we cannot exclude the possibility that biases introduced by misclassification inflated our discrimination. We acknowledge that nonrandom missingness will tend to introduce bias in these estimates. For example, individuals with more severe depression might not pursue postpartum care. Second, while a key strength of this study is the incorporation of academic medical centers and community hospitals and the use of separate development and validation sets, it nonetheless reflects practice patterns in Eastern Massachusetts and Southern New Hampshire in a single health system. Also, the model development and validation was only performed in a subset of all patients within the health system that met the study’s inclusion criteria. Model performance may be different if we had 100% ascertainment of all prepregnancy data in the EHR (i.e., if we did not exclude individuals in which their prenatal care could not be observed) or if we had completed prenatal EPDS score information. Further work will be required to demonstrate model performance in other regions and populations.

In aggregate, this work demonstrates the feasibility of stratifying risk for PPD using electronic health record data routinely collected before discharge after an individual’s delivery. By identifying groups of individuals at greater risk, implementing these models may allow the application of interventions aimed at prevention or more intensive screening that may otherwise be unnecessary or infeasible in the entire clinical population. At a minimum, this model may serve as a baseline that can be augmented by additional screening efforts or biomarkers as they are identified. The next steps of this work involve translating this model into clinical practice and studying how it can be effectively and appropriately used by patients and clinicians to reduce the incidence, severity, and subsequent consequences of PPD.

## Data Availability

Data used in this study has not been made publicly available.

## Acknowledgements

This study was supported by the National Institute of Mental Health (RF1MH132336, Dr. Perlis and Dr. Edlow; R01 MH116270, Dr. Perlis; U54 MH118919, Dr. Edlow), the National Institute of Child Health and Human Development (R01 HD100022-02S2, Dr. Edlow), and Simons Foundation SFARI grant 870754 (Dr. Edlow). The sponsors did not contribute, any aspect of the design and conduct of the study; collection, management, analysis, and interpretation of the data; preparation, review, or approval of the manuscript; and decision, submit the manuscript for publication. The authors had the final responsibility for the decision, submit for publication. Dr. Perlis had full access, all the data in the study and takes responsibility for the integrity of the data and the accuracy of the data analysis.

