## Supplement for "Stratifying Risk for Postpartum Depression at Time of Hospital Discharge"

**Table S1.** Postpartum Depression ICD-10-CM Diagnosis Codes

| **Diagnosis group** | **ICD-10-CM code** | **ICD-10-CM code description** |
|---|---|---|
| Bipolar disorder (CCSR MBD003) | F31.9 | Bipolar disorder, unspecified |
| Bipolar disorder (CCSR MBD003) | F31.81 | Bipolar II disorder |
| Bipolar disorder (CCSR MBD003) | F31.32 | Bipolar disorder, current episode depressed, moderate |
| Bipolar disorder (CCSR MBD003) | F31.75 | Bipolar disorder, in partial remission, most recent episode depressed |
| Bipolar disorder (CCSR MBD003) | F31.30 | Bipolar disorder, current episode depressed, mild or moderate severity, unspecified |
| Bipolar disorder (CCSR MBD003) | F31.31 | Bipolar disorder, current episode depressed, mild |
| Bipolar disorder (CCSR MBD003) | F31.89 | Other bipolar disorder |
| Bipolar disorder (CCSR MBD003) | F31.0 | Bipolar disorder, current episode hypomanic |
| Bipolar disorder (CCSR MBD003) | F31.10 | Bipolar disorder, current episode manic without psychotic features, unspecified |
| Bipolar disorder (CCSR MBD003) | F31.2 | Bipolar disorder, current episode manic severe with psychotic features |
| Bipolar disorder (CCSR MBD003) | F31.62 | Bipolar disorder, current episode mixed, moderate |
| Bipolar disorder (CCSR MBD003) | F31.60 | Bipolar disorder, current episode mixed, unspecified |
| Bipolar disorder (CCSR MBD003) | F31.61 | Bipolar disorder, current episode mixed, mild |
| Bipolar disorder (CCSR MBD003) | F31.77 | Bipolar disorder, in partial remission, most recent episode mixed |
| Bipolar disorder (CCSR MBD003) | F31.71 | Bipolar disorder, in partial remission, most recent episode hypomanic |
| Bipolar disorder (CCSR MBD003) | F31.73 | Bipolar disorder, in partial remission, most recent episode manic |
| Bipolar disorder (CCSR MBD003) | F31.5 | Bipolar disorder, current episode depressed, severe, with psychotic features |
| Bipolar disorder (CCSR MBD003) | F31.4 | Bipolar disorder, current episode depressed, severe, without psychotic features |
| Bipolar disorder (CCSR MBD003) | F31.11 | Bipolar disorder, current episode manic without psychotic features, mild |
| Bipolar disorder (CCSR MBD003) | F31.12 | Bipolar disorder, current episode manic without psychotic features, moderate |
| Bipolar disorder (CCSR MBD003) | F31.64 | Bipolar disorder, current episode mixed, severe, with psychotic features |
| Bipolar disorder (CCSR MBD003) | F34.0 | Cyclothymic disorder |
| Bipolar disorder (CCSR MBD003) | F30.9 | Manic episode, unspecified |
| Bipolar disorder (CCSR MBD003) | F31.13 | Bipolar disorder, current episode manic without psychotic features, severe |
| Bipolar disorder (CCSR MBD003) | F30.8 | Other manic episodes |
| Bipolar disorder (CCSR MBD003) | F30.10 | Manic episode without psychotic symptoms, unspecified |
| Bipolar disorder (CCSR MBD003) | F30.2 | Manic episode, severe with psychotic symptoms |
| Bipolar disorder (CCSR MBD003) | F31.63 | Bipolar disorder, current episode mixed, severe, without psychotic features |
| Bipolar disorder (CCSR MBD003) | F06.33 | Mood disorder due to known physiological condition with manic features |
| Bipolar disorder (CCSR MBD003) | F06.34 | Mood disorder due to known physiological condition with mixed features |
| Depression (CCSR MBD002) | F32.9 | Major depressive disorder, single episode, unspecified |
| Depression (CCSR MBD002) | F53.0 | Postpartum depression |
| Depression (CCSR MBD002) | F32.A | Depression, unspecified |
| Depression (CCSR MBD002) | F33.1 | Major depressive disorder, recurrent, moderate |
| Depression (CCSR MBD002) | F33.41 | Major depressive disorder, recurrent, in partial remission |
| Depression (CCSR MBD002) | F33.9 | Major depressive disorder, recurrent, unspecified |
| Depression (CCSR MBD002) | O90.6 | Postpartum mood disturbance |
| Depression (CCSR MBD002) | F33.0 | Major depressive disorder, recurrent, mild |
| Depression (CCSR MBD002) | F32.1 | Major depressive disorder, single episode, moderate |
| Depression (CCSR MBD002) | F33.2 | Major depressive disorder, recurrent severe without psychotic features |
| Depression (CCSR MBD002) | F32.0 | Major depressive disorder, single episode, mild |
| Depression (CCSR MBD002) | F32.89 | Other specified depressive episodes |
| Depression (CCSR MBD002) | F34.1 | Dysthymic disorder |
| Depression (CCSR MBD002) | F32.4 | Major depressive disorder, single episode, in partial remission |
| Depression (CCSR MBD002) | F32.2 | Major depressive disorder, single episode, severe without psychotic features |
| Depression (CCSR MBD002) | F32.81 | Premenstrual dysphoric disorder |
| Depression (CCSR MBD002) | F33.3 | Major depressive disorder, recurrent, severe with psychotic symptoms |
| Depression (CCSR MBD002) | F32.3 | Major depressive disorder, single episode, severe with psychotic features |
| Depression (CCSR MBD002) | F33.8 | Other recurrent depressive disorders |
| Depression (CCSR MBD002) | F06.31 | Mood disorder due to known physiological condition with depressive features |
| Depression (CCSR MBD002) | F06.32 | Mood disorder due to known physiological condition with major depressive-like episode |

| Diagnosis group | ICD-10-CM code | ICD-10-CM code description |
|---|---|---|
| Depression (CCSR MBD002) | F32.8 | Other depressive episodes |
| Depression (CCSR MBD002) | F06.34 | Mood disorder due to known physiological condition with mixed features |
| Schizophrenia (CCSR MBD001) | F53 | Puerperal psychosis |
| Schizophrenia (CCSR MBD001) | F29 | Unspecified psychosis not due to a substance or known physiological condition |
| Schizophrenia (CCSR MBD001) | F53.1 | Puerperal psychosis |
| Schizophrenia (CCSR MBD001) | F20.9 | Schizophrenia, unspecified |
| Schizophrenia (CCSR MBD001) | F25.9 | Schizoaffective disorder, unspecified |
| Schizophrenia (CCSR MBD001) | F22 | Delusional disorders |
| Schizophrenia (CCSR MBD001) | F25.0 | Schizoaffective disorder, bipolar type |
| Schizophrenia (CCSR MBD001) | F23 | Brief psychotic disorder |
| Schizophrenia (CCSR MBD001) | F06.1 | Catatonic disorder due to known physiological condition |
| Schizophrenia (CCSR MBD001) | F20.0 | Paranoid schizophrenia |
| Schizophrenia (CCSR MBD001) | F14.159 | Cocaine abuse with cocaine-induced psychotic disorder, unspecified |
| Schizophrenia (CCSR MBD001) | F20.89 | Other schizophrenia |
| Schizophrenia (CCSR MBD001) | F28 | Other psychotic disorder not due to a substance or known physiological condition |
| Schizophrenia (CCSR MBD001) | F06.0 | Psychotic disorder with hallucinations due to known physiological condition |
| Schizophrenia (CCSR MBD001) | F12.951 | Cannabis use, unspecified with psychotic disorder with hallucinations |
| Schizophrenia (CCSR MBD001) | F12.959 | Cannabis use, unspecified with psychotic disorder, unspecified |
| Schizophrenia (CCSR MBD001) | F19.959 | Other psychoactive substance use, unspecified with psychoactive substance-induced psychotic disorder, unspecified |
| Schizophrenia (CCSR MBD001) | F20.1 | Disorganized schizophrenia |
| Schizophrenia (CCSR MBD001) | F20.2 | Catatonic schizophrenia |
| Schizophrenia (CCSR MBD001) | F20.81 | Schizophreniform disorder |
| Schizophrenia (CCSR MBD001) | F25.1 | Schizoaffective disorder, depressive type |
| Schizophrenia (CCSR MBD001) | F25.8 | Other schizoaffective disorders |
| Schizophrenia (CCSR MBD001) | F19.951 | Other psychoactive substance use, unspecified with psychoactive substance-induced psychotic disorder with hallucinations |
| Schizophrenia (CCSR MBD001) | F20.3 | Undifferentiated schizophrenia |
| Schizophrenia (CCSR MBD001) | F10.959 | Alcohol use, unspecified with alcohol-induced psychotic disorder, unspecified |
| Schizophrenia (CCSR MBD001) | F12.259 | Cannabis dependence with psychotic disorder, unspecified |
| Schizophrenia (CCSR MBD001) | F14.951 | Cocaine use, unspecified with cocaine-induced psychotic disorder with hallucinations |
| Schizophrenia (CCSR MBD001) | F15.159 | Other stimulant abuse with stimulant-induced psychotic disorder, unspecified |
| Schizophrenia (CCSR MBD001) | F15.950 | Other stimulant use, unspecified with stimulant-induced psychotic disorder with delusions |
| Schizophrenia (CCSR MBD001) | F21 | Schizotypal disorder |
| Suicidality (CCSR MBD012) | R45.851 | Suicidal ideations |
| Suicidality (CCSR MBD012) | T14.91XA | Suicide attempt, initial encounter |
| Suicidality (CCSR MBD012) | T50.902A | Poisoning by unspecified drugs, medicaments and biological substances, intentional self-harm, initial encounter |
| Suicidality (CCSR MBD012) | T39.312A | Poisoning by propionic acid derivatives, intentional self-harm, initial encounter |
| Suicidality (CCSR MBD012) | T39.1X2A | Poisoning by 4-Aminophenol derivatives, intentional self-harm, initial encounter |
| Suicidality (CCSR MBD012) | T50.992A | Poisoning by other drugs, medicaments and biological substances, intentional self-harm, initial encounter |
| Suicidality (CCSR MBD012) | X83.8XXA | Intentional self-harm by other specified means, initial encounter |
| Suicidality (CCSR MBD012) | T42.6X2A | Poisoning by other antiepileptic and sedative-hypnotic drugs, intentional self-harm, initial encounter |
| Suicidality (CCSR MBD012) | T43.222A | Poisoning by selective serotonin reuptake inhibitors, intentional self-harm, initial encounter |
| Suicidality (CCSR MBD012) | T43.592A | Poisoning by other antipsychotics and neuroleptics, intentional self-harm, initial encounter |
| Suicidality (CCSR MBD012) | T54.92XA | Toxic effect of unspecified corrosive substance, intentional self-harm, initial encounter |
| Suicidality (CCSR MBD012) | X78.9XXA | Intentional self-harm by unspecified sharp object, initial encounter |
| Suicidality (CCSR MBD012) | T14.91 | Suicide attempt |
| Suicidality (CCSR MBD012) | T40.4X2A | Poisoning by other synthetic narcotics, intentional self-harm, initial encounter |
| Suicidality (CCSR MBD012) | T51.0X2A | Toxic effect of ethanol, intentional self-harm, initial encounter |
| Suicidality (CCSR MBD012) | X78.1XXA | Intentional self-harm by knife, initial encounter |
| Suicidality (CCSR MBD012) | T39.012A | Poisoning by aspirin, intentional self-harm, initial encounter |
| Suicidality (CCSR MBD012) | T39.92XA | Poisoning by unspecified nonopioid analgesic, antipyretic and antirheumatic, intentional self-harm, initial encounter |
| Suicidality (CCSR MBD012) | T42.4X2A | Poisoning by benzodiazepines, intentional self-harm, initial encounter |

| Diagnosis group | ICD-10-CM code | ICD-10-CM code description |
|---|---|---|
| Suicidality (CCSR MBD012) | T42.8X2A | Poisoning by antiparkinsonism drugs and other central muscle-tone depressants, intentional self-harm, initial encounter |
| Suicidality (CCSR MBD012) | T43.292A | Poisoning by other antidepressants, intentional self-harm, initial encounter |
| Suicidality (CCSR MBD012) | T44.7X2A | Poisoning by beta-adrenoreceptor antagonists, intentional self-harm, initial encounter |
| Suicidality (CCSR MBD012) | T45.2X2A | Poisoning by vitamins, intentional self-harm, initial encounter |
| Suicidality (CCSR MBD012) | T50.912A | Poisoning by multiple unspecified drugs, medicaments and biological substances, intentional self-harm, initial encounter |
| Suicidality (CCSR MBD012) | T65.92XA | Toxic effect of unspecified substance, intentional self-harm, initial encounter |
| Suicidality (CCSR MBD012) | X78.8XXA | Intentional self-harm by other sharp object, initial encounter |
| Postpartum depression | F53.0 | Postpartum depression |
| Postpartum depression | F53 | Puerperal psychosis |
| Postpartum depression | O90.6 | Postpartum mood disturbance |
| Postpartum depression | F53.1 | Puerperal psychosis |

**Table S2**. Postpartum Depression Medications

| Drug group | Drug ingredient |
|---|---|
| Antidepressant | sertraline HCl |
| Antidepressant | escitalopram oxalate |
| Antidepressant | fluoxetine HCl |
| Antidepressant | citalopram hydrobromide |
| Antidepressant | bupropion HCl |
| Antidepressant | venlafaxine HCl |
| Antidepressant | duloxetine HCl |
| Antidepressant | amitriptyline HCl |
| Antidepressant | nortriptyline HCl |
| Antidepressant | mirtazapine |
| Antidepressant | paroxetine HCl |
| Antidepressant | desvenlafaxine succinate |
| Antidepressant | fluvoxamine maleate |
| Antidepressant | clomipramine HCl |
| Antidepressant | vilazodone HCl |
| Antidepressant | vortioxetine hydrobromide |
| Antidepressant | desipramine HCl |
| Antidepressant | desvenlafaxine |
| Antidepressant | imipramine HCl |
| Antipsychotic | quetiapine fumarate |
| Antipsychotic | olanzapine |
| Antipsychotic | risperidone |
| Antipsychotic | lurasidone HCl |
| Antipsychotic | haloperidol |
| Antipsychotic | perphenazine |
| Antipsychotic | ziprasidone HCl |
| Antipsychotic | chlorpromazine HCl |
| Antipsychotic | brexpiprazole |
| Antipsychotic | aripiprazole lauroxil |
| Antipsychotic | haloperidol decanoate |
| Antipsychotic | loxapine succinate |
| Antipsychotic | clozapine |
| Antipsychotic | paliperidone |
| Bipolar medication | lamotrigine |
| Bipolar medication | aripiprazole |
| Bipolar medication | lithium carbonate |
| Bipolar medication | carbamazepine |
| Bipolar medication | divalproex sodium |
| Bipolar medication | eslicarbazepine acetate |
| Bipolar medication | valproic acid |

**Table S3.** Sociodemographic and clinical characteristics in mothers with and without post-partum depression within 6-months of delivery in the full study cohort.

| **Characteristic** | **PPD**, N = 2,703 | **No PPD**, N = 26,465 | **Difference (95% CI)**[1,2] |
|---|---|---|---|
| Age at delivery (y), Median (IQR) | 33 (30 – 36) | 33 (30 – 36) | -0.35 (-0.54 to -0.16) |
| Race, n (%) | | | 0.15 (0.11 to 0.19) |
| Asian | 240 (9.0) | 3,435 (13) | |
| Black | 178 (6.7) | 1,818 (7.0) | |
| Two or more | 50 (1.9) | 430 (1.7) | |
| White | 1,988 (75) | 18,104 (70) | |
| Other | 196 (7.4) | 2,195 (8.4) | |
| Unknown | 51 | 483 | |
| Ethnicity, n (%) | | | 0.02 (-0.02 to 0.06) |
| Hispanic | 313 (12) | 2,850 (11) | |
| Non-Hispanic | 2,319 (88) | 22,765 (89) | |
| Unknown | 71 | 850 | |
| Marital status, single, n (%) | 584 (22) | 4,380 (17) | 5.1% (3.4% to 6.7%) |
| Public insurance, n (%) | 361 (13) | 3,237 (12) | 1.1% (-0.24% to 2.5%) |
| Education, college degree, n (%) | 1,826 (68) | 19,329 (73) | -5.5% (-7.3% to -3.6%) |
| Employment, full-time, n (%) | 1,803 (67) | 17,637 (67) | 0.06% (-1.8% to 1.9%) |
| Non-English primary language, n (%) | 128 (4.7) | 1,821 (6.9) | -2.1% (-3.0% to -1.3%) |
| Pre-pregnancy BMI, Median (IQR) | 25 (22 – 30) | 24 (21 – 27) | 1.5 (1.2 to 1.8) |
| Unknown | 557 | 5,657 | |
| Gestational diabetes, n (%) | 546 (20) | 4,732 (18) | 2.3% (0.72% to 3.9%) |
| Preeclampsia, n (%) | 465 (17) | 3,332 (13) | 4.6% (3.1% to 6.1%) |
| Prenatal visit index, Median (IQR) | 11 (9 – 13) | 11 (9 – 12) | 0.10 (-0.03 to 0.23) |
| Prenatal EPDS, Median (IQR) | 6 (3 – 9) | 3 (1 – 6) | 2.6 (2.4 to 2.7) |
| Delivery LOS (days), Median (IQR) | 3.06 (2.50 – 4.14) | 2.85 (2.27 – 3.75) | 0.62 (0.46 to 0.78) |
| Multiple birth, n (%) | 99 (3.7) | 823 (3.1) | 0.55% (-0.21% to 1.3%) |
| Pre-term delivery, n (%) | 370 (14) | 2,505 (9.5) | 4.2% (2.9% to 5.6%) |
| Postpartum care, n (%) | 2,326 (86) | 21,608 (82) | 4.4% (3.0% to 5.8%) |
| Postpartum EPDS >= 13, n (%) | 692 (26) | 0 (0) | 26% (24% to 27%) |

*EPDS: Edinburgh Postpartum depression score (EPDS), LOS: Length of stay;*
[1] *Welch Two Sample t-test; Standardized Mean Difference; Two sample test for equality of proportions;*
[2] *CI = Confidence Interval*

**Table S4.** Model discrimination and calibration by population subgroups

| Subgroup | Value | AUROC | 95% CI LL | 95% CI UL | Brier Class | 95% CI LL | 95% CI UL |
|---|---|---|---|---|---|---|---|
| Age | <30 | 0.723 | 0.690 | 0.755 | 0.095 | 0.085 | 0.104 |
| | 30-40 | 0.719 | 0.704 | 0.735 | 0.086 | 0.081 | 0.090 |
| | >40 | 0.742 | 0.678 | 0.798 | 0.092 | 0.073 | 0.111 |
| Race | White | 0.718 | 0.701 | 0.733 | 0.093 | 0.088 | 0.098 |
| | Black | 0.774 | 0.726 | 0.815 | 0.078 | 0.065 | 0.090 |
| | Other | 0.711 | 0.656 | 0.766 | 0.078 | 0.064 | 0.092 |
| | Asian | 0.724 | 0.676 | 0.773 | 0.061 | 0.051 | 0.070 |
| | Two or more | 0.723 | 0.624 | 0.827 | 0.123 | 0.087 | 0.160 |
| Ethnicity | Non-Hispanic | 0.717 | 0.702 | 0.732 | 0.088 | 0.084 | 0.092 |
| | Hispanic | 0.729 | 0.689 | 0.767 | 0.089 | 0.078 | 0.102 |
| Hospital Type | Academic medical center | 0.709 | 0.691 | 0.728 | 0.092 | 0.087 | 0.097 |
| | Community hospital | 0.742 | 0.720 | 0.764 | 0.081 | 0.075 | 0.087 |

**Figure S1.** Feature importance plot in the primary model

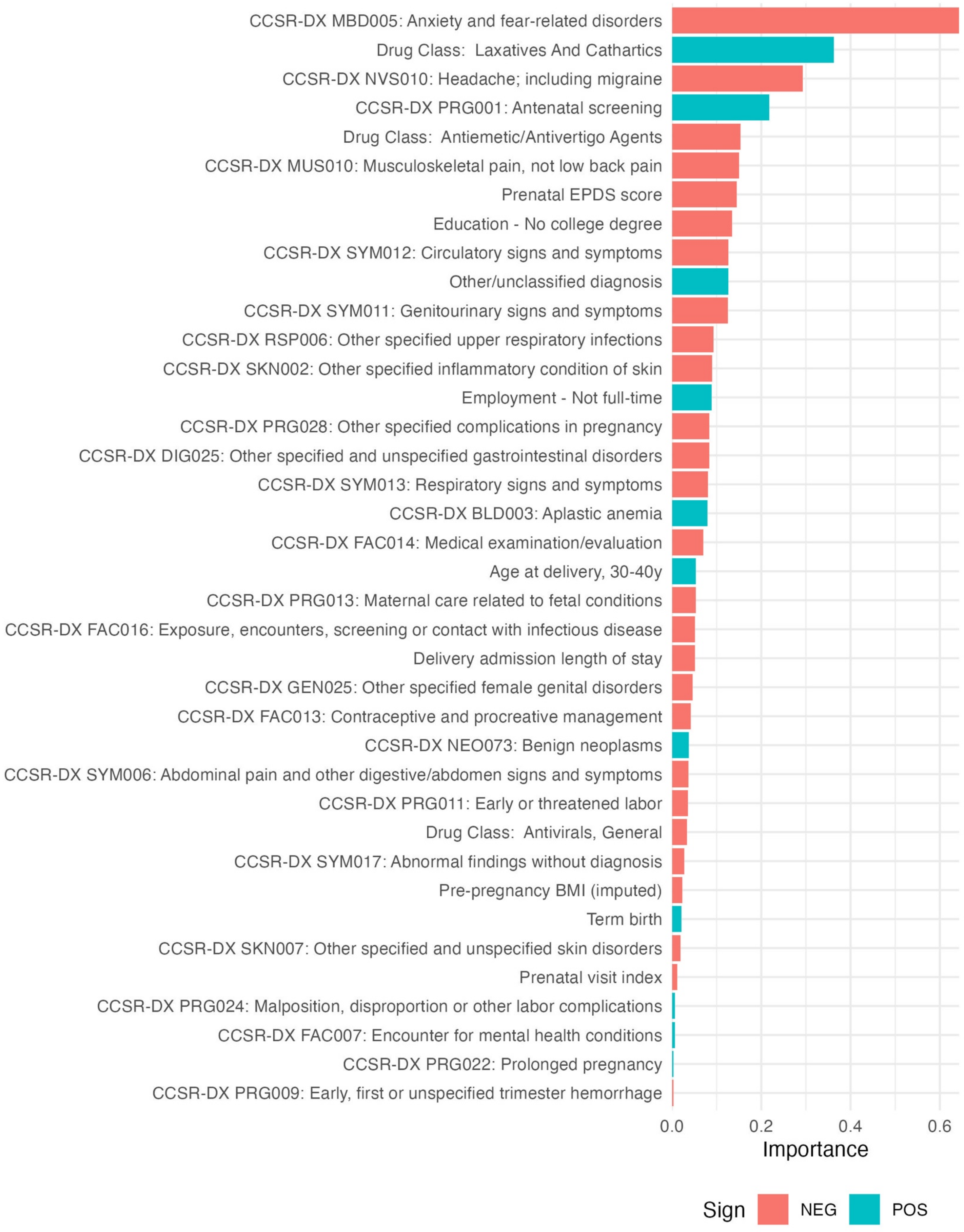
